# Smartphone Imaging for Remote Monitoring of Inflammatory Arthritis in a Real-World Cohort: Longitudinal Evaluation of a Machine Learning-Based Finger Fold Biomarker

**DOI:** 10.64898/2026.07.30.26359295

**Authors:** Cinja Nadana Koller, Jules Maglione, Marc Blanchard, Alexandre Dumusc, Diana Dan, Laure Brulhart, Michael Nissen, Michael Andor, Raphael Micheroli, Almut Scherer, Christos Polysopoulos, Andrea Rubbert-Roth, Christof Iking-Konert, Tobias Manigold, Burkhard Möller, Chrisa Manolarki, Jeroen Geurts, Thomas Hügle

**Affiliations:** Department of Rheumatology, University Hospital Lausanne (CHUV) and University of Lausanne (UNIL), Lausanne, Switzerland; Department of Rheumatology, Réseau Hospitalier Neuchâteloise de la Chaux de fonds, Switzerland; Department of Rheumatology, University Hospital of Geneva (HUG) and University of Geneva, Geneva, Switzerland; Rheumatologie im Zürcher Oberland (RZO), Uster, Switzerland; Department of Rheumatology, University Hospital of Zurich (USZ) and University of Zurich (UZH), Zurich, Switzerland; SCQM Foundation, Zurich, Switzerland; Department of Rheumatology, Health Ostschweiz, Kantonsspital St. Gallen (HOCH), Switzerland; Department of Rheumatology, Stadtspital Zurich, Zurich, Switzerland; Rheumatology Inselspital, Bern, Switzerland; Rheumabasel, Basel, Switzerland

## Abstract

**Background:** Smartphone enabled remote patient monitoring has the potential to complement conventional follow-up in inflammatory arthritis. We previously presented the finger fold index (FFI) derived from hand photographs as ratio of automated detected proximal interphalangeal (PIP) joint diameter and surface of dorsal finger folds as a digital biomarker for clinical joint swelling and disease activity in rheumatoid arthritis (RA) and psoriatic arthritis (PsA).

**Objective:** To evaluate the feasibility, image quality, patient engagement, and clinical utility of both HCP- and patient-collected hand photographs integrated into a national rheumatology registry, and to assess the performance of the FFI as an image-derived digital biomarker for clinical joint swelling of the proximal interphalangeal joints in a real-world arthritis cohort.

**Methods:** In this longitudinal multicenter study, a photo function with written instructions were integrated into the Swiss Clinical Quality Management in Rheumatic Diseases (SCQM) registry and their mySCQM mobile application, respectively. Patients with RA or PsA contributed longitudinal smartphone photographs together with patient-reported outcomes (PROs) via the mySCQM mobile application while health care professionals (HCPs) acquired images during routine visits. After manual quality assessment, images were processed using an automated computer vision pipeline to derive the FFI, a digital biomarker based on dorsal finger-fold morphology. Image quality was evaluated for both HCP- and participant-collected photographs, and patient engagement was assessed. Associations between FFI, clinical proximal interphalangeal (PIP) joint swelling, RADAI-5, DAS28-CRP and longitudinal changes were assessed. A generalized linear mixed model was used to estimate the association between FFI and joint swelling while accounting for repeated measures and within-subject correlations.

**Results:** Between 2023 and 2025, 374 RA and PsA patients were included. HCPs captured 977 hand images while 174 patients collected 1228 hand images via the mySCQM app. Patients demonstrated sustained engagement after instruction, contributing a mean of seven images during data collection. Following quality control, 1729 hand images comprising 4048 PIP joints were included for analysis. Image quality was comparable between patient-acquired and HCP-acquired photographs; 78.3% of the patient-acquired vs. 73.5% of the HCP-acquired hand images were suitable to run the ML-model. 23.1% of the cropped joints had to be removed after the running of the FFI algorithm due to false diameter or finger fold detection e.g. due to wrong hand positioning. In images taken by HCPs, mean FFI and DAS28-CRP were weakly but significantly correlated (Spearman’s ρ = 0.164; 95% CI [0.004 to 0.317]; p = 0.039). Conversely, RADAI-5 scores did not correlate with the mean FFI in RA patients (r = 0.007, *p* = 0.932, 95% CI [-0.169-0.183]). At follow-up visits, clinical swelling resolved in 40 joints, of which in 68.0% the direction of the delta FFI was concordant with the clinical change. In contrast, 13 joints developed incident clinical swelling, of which 87.5% had a direction of the delta FFI that was concordant with the clinical change. However, the GLMM showed no significant associations between swelling and joint location or time-varying FFI, and no evidence of interaction between FFI and PIP joint.

**Conclusion:** Integration of patient self-imaging into a remote monitoring application for inflammatory arthritis is feasible and achieves image quality comparable to clinician acquired photographs. The FFI derived from collected images shows association with clinical joint swelling and disease activity scores, but not PROs. In a substantial proportion of images, the FFI algorithm could not be applied because of insufficient image quality. More standardized image acquisition and further refinement of the FFI algorithm are warranted.

## Introduction

In inflammatory arthritis, monitoring disease activity is essential to enable treatment adjustment and treat-to-target strategies as persistent inflammation may lead to irreversible joint damage [1], [2], [3]. In routine clinical practice, however, disease activity is primarily assessed during scheduled outpatient visits, which are often separated by several months. Between visits, patient well-being and disease activity are commonly monitored using patient-reported outcomes (PROs) [4]. Although objective measures such as C-reactive protein (CRP), musculoskeletal ultrasound and clinical joint examination provide greater specificity for inflammation, their integration into remote patient monitoring remains limited due to logistical constraints, including the need for specialized equipment and trained personnel [5], [6], [7]. Furthermore, PROs and systemic inflammatory biomarkers alone have limited specificity for inflammatory disease activity, as symptoms may be influenced by fibromyalgia, osteoarthritis, psychosocial factors, or other comorbidities [8], [9]. Consequently, additional objective digital biomarkers are needed to complement PRO-based remote monitoring. Wearables and patient-acquired smartphone imaging have emerged as promising modalities for this purpose [8], [9], [10], [11], [12], [13].

Self-imaging of the hands and other joints has gained increasing interest in inflammatory arthritis, particularly in rheumatoid arthritis, where hand and wrist involvement is highly prevalent, and in psoriatic arthritis, in which peripheral manifestations such as arthritis, dactylitis, and nail disease can also be captured to complement patient-reported outcomes. These approaches may enable scalable remote assessment of joint status and provide a form of asynchronous, patient-acquired joint evaluation. This concept has been implemented at scale in our work within a national registry, supporting the feasibility of remote imaging in routine care and enabling additional clinical assessment in situations such as suspected disease flares identified together with PROs.

Among the clinical manifestations of inflammatory arthritis, joint swelling is one of the most objective indicators of active inflammation and remains a central component of disease activity assessment. However, reliable assessment of swelling requires clinical expertise and is difficult for patients to perform themselves [14], [15]. Recent advances in artificial intelligence and computer vision have enabled automated extraction of quantitative image features from standard photographs, raising the possibility that visible manifestations of inflammation may be assessed remotely. Smartphone imaging is particularly attractive because it is inexpensive, widely available, and can be repeatedly performed by patients at home, enabling longitudinal documentation of hand morphology between clinic visits.

We previously demonstrated that convolutional neural networks can automatically detect and quantify dorsal finger-fold patterns on hand photographs, resulting in the Finger Fold Index (FFI), an image-derived digital biomarker associated with proximal interphalangeal (PIP) joint swelling in patients with rheumatoid arthritis (RA) and psoriatic arthritis (PsA) [16], [17]. However, these studies were performed under controlled imaging conditions and did not address whether such image-derived biomarkers remain informative in routine clinical practice or when photographs are acquired repeatedly by patients themselves.

A major challenge for deploying digital image biomarkers is not only algorithm performance but also the acquisition of standardized longitudinal image data in real-world settings. Patient-generated photographs vary in lighting, positioning, camera angle, and image quality, all of which may affect downstream image analysis. Whether patients can reliably contribute photographs of sufficient quality for automated processing has not been systematically investigated. Likewise, the feasibility of integrating longitudinal smartphone imaging into established remote patient monitoring workflows remains largely unknown.

To address these questions, we embedded standardized hand-photograph acquisition into the Swiss Clinical Quality Management in Rheumatic Diseases (SCQM) registry and its companion patient application mySCQM. Healthcare professionals captured photographs during routine visits, while patients were encouraged to upload hand photographs longitudinally from home together with their routine PROs. The application served exclusively as a platform for image acquisition and upload, whereas all computer vision analyses were performed offline using our previously developed machine-learning pipeline. We evaluated the feasibility and quality of patient-generated images in comparison with healthcare professional–acquired photographs and investigated whether the Finger Fold Index was associated with clinical joint swelling, disease activity, and longitudinal changes over time in a multicenter real-world cohort of patients with RA and PsA.

## Methods

### Study design

We conducted a multicenter, observational longitudinal study including patients diagnosed with RA or PsA. Participants had to be at least 18 years old, fulfil the ACR/EULAR 2010 RA classification criteria or CASPAR criteria, and have hand involvement. Eligible participants had to be either already included in the Swiss Clinical Quality Management (SCQM) registry in Switzerland or be included before inclusion into the study. Participants were excluded if they had undergone joint arthrodesis or joint replacement of hand joints, or had concomitant microcrystal disease or osteoarthritis of hand joints. Patients with lymphoedema were also excluded. All participants provided written informed consent. The study was approved by the ethical commission CER-VD (2020-00033) and conducted in accordance with the Declaration of Helsinki.

### Data collection

Nine rheumatology departments in Switzerland were trained to capture hand photographs from RA and PsA patients via the integrated photo function in the SCQM registry, using either smartphones or tablets. One center used a photo box to take images. Instructions on how to position the patient’s hands were distributed and were available at all times during the study within the registry in three different languages. Each hand was to be placed separately, flat, with fingers spread on a white background. The photograph was supposed to be taken in a vertical position approximately 20 cm above the dorsal side of the hand. Data collection took place between 2023 and 2025. Photographs were taken during routine rheumatology consultations alongside clinical variables. Clinical variables included DAS28-CRP, CRP (mg/l), Physician Global Assessment, swelling in PIP joints, skin involvement, rheumatoid factor and anti–cyclic citrullinated peptide (anti-CCP) status. Demographic variables and patient characteristics such as age, sex, disease duration, weight or height were obtained from the SCQM registry.

HCPs were instructed to encourage participants to use the mySCQM app (the SCQM registry application for reporting patient-reported outcomes) to regularly take photographs of their hands via the app and RA patients to fill in the RADAI-5 questionnaire, a PRO for disease activity. Participants were asked to take at least one photograph and complete the PROs on the day of the visit to allow comparison of image quality between images taken by HCPs and those taken by patients. No further specific guidance was provided in order to explore how participants would use this new functionality for taking hand photographs. Instructions on how to take photographs were available within the app at all times during the study, and upon first consent, participants were required by the app structure to go through the information and instructions.

**Figure 1.**
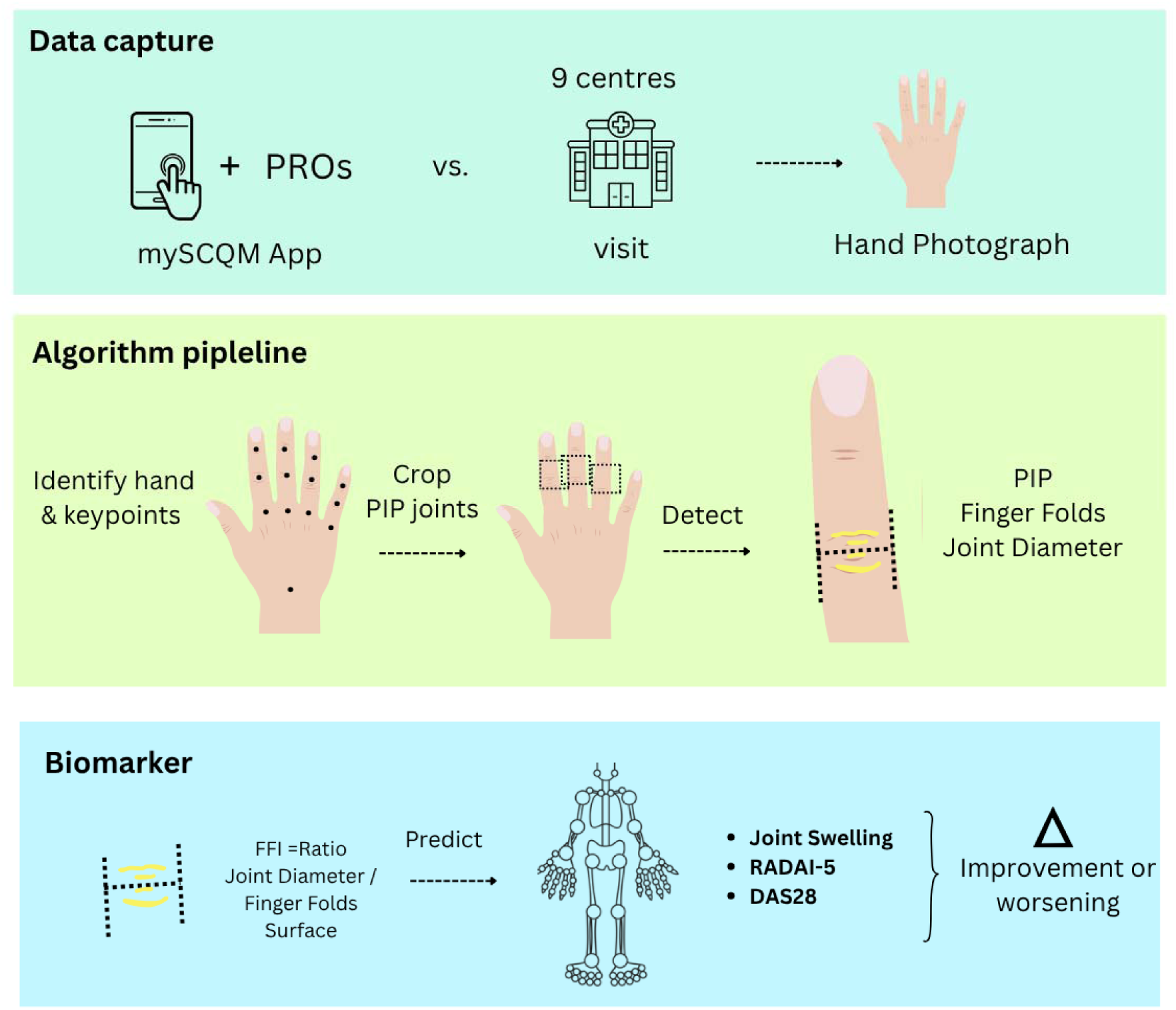
Study design and implementation. After instructions, patients collected hand images via the mySCQM app on their smartphone and provided RADAI-5 scores. Photos were also taken during regular visits along with DAS28-CRP. FFI = Finger Fold Index.

### Image quality check and algorithm processing

Hand photographs were first sorted by quality. Images below 400 kilobytes were removed before being analyzed by a machine learning (ML) pipeline for automated detection and processing of PIP joints. Images that did not show a hand (e.g., feet, knee, or other body parts) were excluded. Palmar-side hand images, as well as laterally acquired images, were not retained. Images with casts or bandages were also removed.

Afterwards, images were processed using MediaPipe and a U-Net model, as previously described by Koller et al. [17]. In short, the MediaPipe model detects landmarks on the hand, including the PIP joints, and crops these joints. Computer vision techniques are then used to identify and calculate the joint diameter. Finally, a U-Net model detects the fold pixels on the joints, allowing for the calculation of the FFI as the ratio of diameter to pixel fold area.

The resulting cropped joint images, including finger folds and diameter visualizations, underwent a second quality check by one person. In this step, single cropped joint images with missing or incorrect diameter or fold annotations were removed, together with their respective FFI values. This included, for example, joints showing fold delineation across two joints, folds located outside the joint area, or features corresponding to hair on the finger rather than skin folds.

### Statistical analysis

Continuous variables were described with mean and standard deviation (SD) or median and interquartile range (IQR) whereas categorical were described with numbers and percentage. The data was analyzed with SPSS Version 29.0.2.0 (20) and statistical significance was set at p < 0.05 with 95% Confidence Intervals (CI). Data was tested for normality using the Shapiro-wilk test. Due to non-normality of the data, Pearson correlation was used to assess correlations. Longitudinal relationships were performed by correlating changes in the DAS28-CRP and mean FFI between the baseline and the first follow-up visit as well as the joint specific swelling and joint specific FFI changes. Associations between the FFI and PIP joint swelling were analyzed using generalized linear mixed models (GLMMs). The GLMM was fitted to examine predictors of swelling and included FFI and PIP joint number (2, 3, 4 for each side) as fixed effects, with an intercept term included. A subject-specific random intercept was included to account for repeated measurements within patients, allowing baseline swelling probability to vary across individuals. Repeated measurements across visits and PIP joints were modelled with a first-order autoregressive covariance structure to account for within-subject correlation over time and across repeated measurements (FFI).

## Results

### Image collection

Of the 2205 collected images, HCPs collected 977 images of 374 participants while 174 participants self-collected 1228 images. After the two rounds of quality review, 719 images (73.6%) of 209 participants collected by HCPs and 961 images (78.3%) from 159 participants that were self-collected were included in the analysis (Fig. 2).

**Figure 2.**
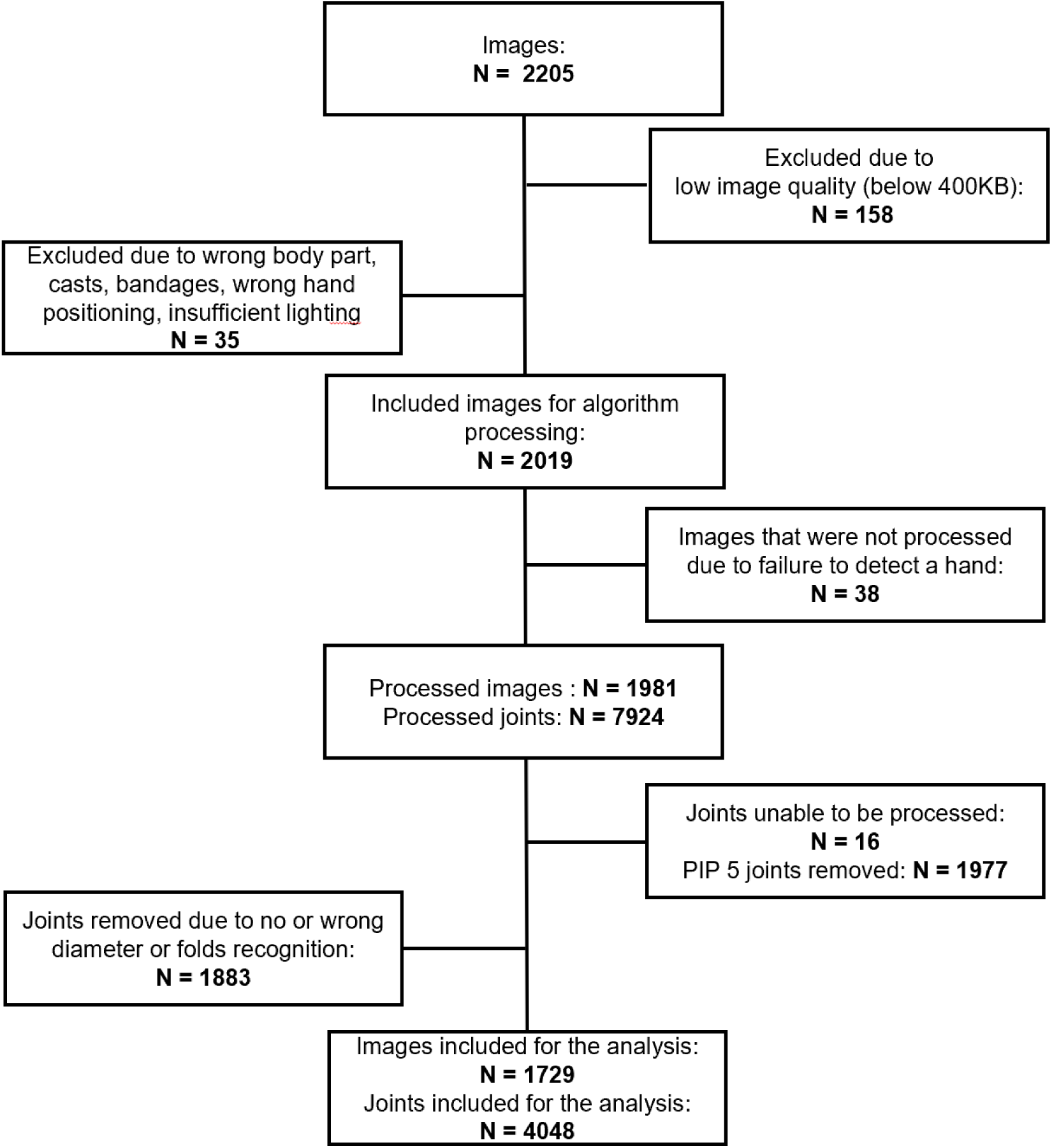
Hand image and joint image inclusion flow.

During the second round of quality review, which assessed individual processed joints of images from both HCPs and participant-collected images, 23.13% of processed joints had to be excluded. The main reasons were missing or incorrect detection of diameters or folds, most commonly due to insufficient finger spreading. In some cases, jewellery or hair were incorrectly identified as finger folds (Fig. 3). Minor quality issues were accepted.

**Figure 3.**
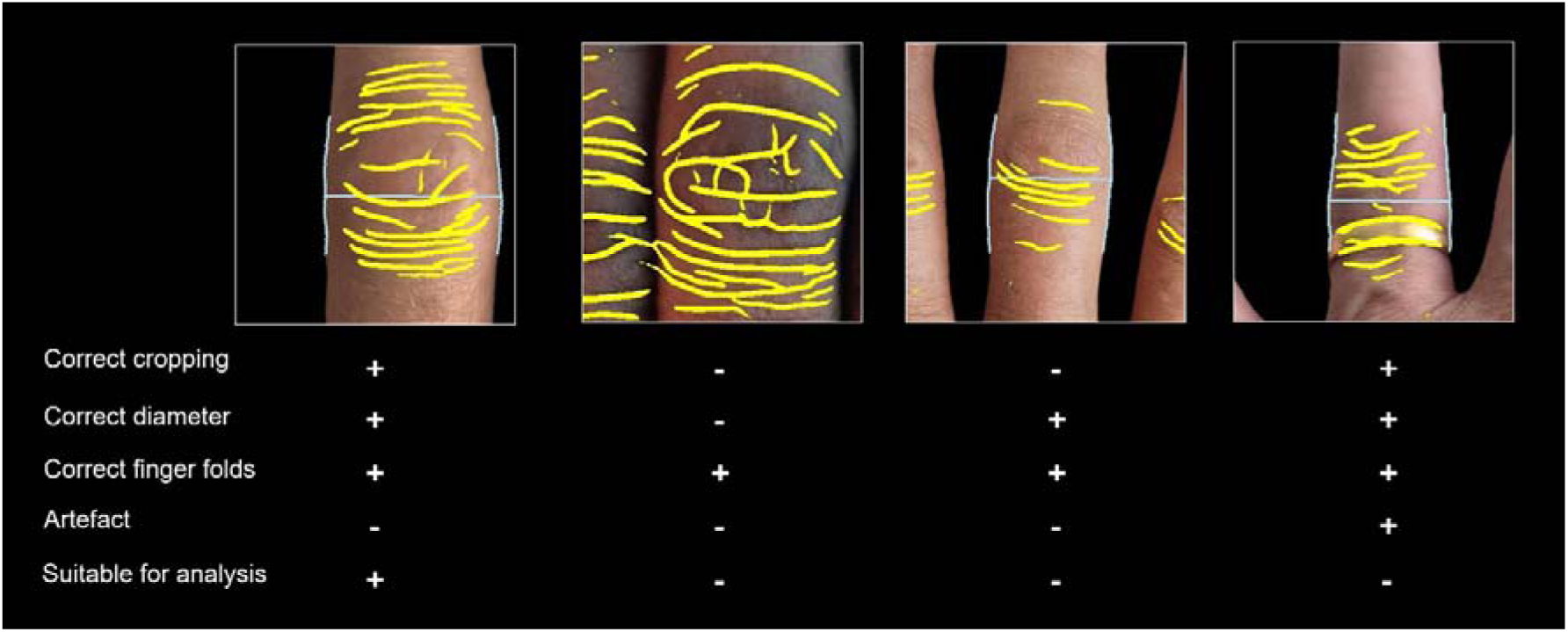
Image quality assessment for automated diameter and dorsal finger fold detection on cropped Proximal Interphalangeal (PIP) joints. Representative examples of images acquired with smartphone cameras by patients or healthcare professionals, illustrating the quality criteria for automated analysis: correct finger diameter, correct visualization of finger folds, and absence of artefacts. Only images meeting all criteria were considered suitable for analysis. Yellow lines indicate the automatically detected finger fold patterns.

Participants were predominantly female 71% (n = 148). The majority were diagnosed with RA 79% (n = 165), while 52% (n = 108) were anti-CCP positive and 55% (n = 115) were RF positive (Table 1). In total, 378 study visits were recorded, with participants contributing data and images from 1 to 6 visits. Median time to follow-up (visit 1) after baseline was 184 days with an IQR of 209.25 days. The RA and PsA cohorts differed with respect to sex, age and rheumatoid factor (RF) status but were otherwise comparable.

**Table 1.** Patient characteristics at baseline visit.

| Variable | Total N = 209 (100) | RA N = 165 (78.9) | PsA N = 44 (21.1) | P-value |
| --- | --- | --- | --- | --- |
| Age, mean (SD) | 60.88 (13.316) | 62.15 (13.145) | 56.09 (13.002) | **0.007** |
| Female, n (%) | 148 (70.8) | 131 (79.4) | 17 (38.6) | **<0.001** |
| BMI, median, (IQR) | 25.2832 (6.35)<br>Missing N = 133 | 24.5448 (6.32)<br>Missing N = 107 | 26.5400 (6.03)<br>Missing N = 26 | 0.153 |
| Disease duration, median (IQR) | 13 (13)<br>Missing N = 134 | 14 (14)<br>Missing N = 9 | 12 (15)<br>Missing N = 3 | 0.216 |
| Positive Anti-ccp, n (%) | 108 (51.7)<br>Missing N = 54 | 108 (65.5)<br>Missing N = 11 | 0 (0)<br>Missing N = 43 | 0.303 |
| Positive RF, n (%) | 115 (55.0)<br>Missing N = 20 | 113 (68.5)<br>Missing N = 9 | 2 (4.5)<br>Missing N = 11 | **<0.001** |
| DAS28, median (IQR) | 2.3 (2)<br>Missing N = 70 | 2.4 (2)<br>Missing N = 48 | 1.7 (1.4)<br>Missing N = 22 | 0.065 |

*Normally distributed variables are described with mean (SD), non-normally distributed variables with median (IQR).*

### Healthcare professional collected image analyses

#### DAS28-CRP and association with the mean FFI

Mean FFI showed a weak positive correlation with DAS28-CRP in the overall cohort (Spearman’s ρ = 0.164; 95% CI [0.004 to 0.317]; p = 0.039, N=158) which was statistically significant. The correlation did not remain significant for the RA (Spearman’s ρ = 0.163; 95% CI [−0.007 to 0.323]; p = 0.052, N=143) and the PsA cohort separate (Spearman’s ρ = 0.343; 95% CI [−0.221 to 0.735]; p = 0.211, N=15). Changes in mean FFI between visit 1 and 2 in the mixed cohort were not correlated with changes in DAS28-CRP (Spearman’s ρ = −0.002; 95% CI [−0.434 to 0.431]; p = 0.994, N=22).

#### Association of FFI with PIP joint swelling

The GLMM did not identify the time-varying FFI as a statistically significant predictor (Table 2). The random intercept showed significant between-subject variability in swelling (variance = 5.111, p < .001).

**Table 2.** Fixed-Effect Coefficients, Odds Ratios, and 95% Confidence Intervals from the GLMM for PIP joint swelling.

| **Model Terms**<br>**(Predictors)** | **B (Coefficient)** | **Std. Error** | ***p* value** | **Exp(B)** | **95% CI for Exp(B)** |
| --- | --- | --- | --- | --- | --- |
| Intercept | 5.319 | 1.0711 | <0.001 | 204.198 | 24.965 – 1670.190 |
| Mean FFI | -0.056 | 0.0185 | **0.002** | 0.945 | 0.912 – 0.980 |
| FFI | 0.009 | 0.0140 | 0.519 | 1.009 | 0.982 – 1.037 |
| PIP 2 Right | -0.948 | 0.5067 | 0.061 | 0.387 | 0.143 – 1.047 |
| PIP 3 Right | -1.154 | 0.5536 | **0.037** | 0.315 | 0.106 – 0.934 |
| PIP 4 Right | -0.644 | 0.5033 | 0.201 | 0.525 | 0.196 – 1.409 |
| PIP 2 Left | 0.734 | 0.5234 | 0.161 | 2.083 | 0.746 – 5.816 |
| PIP 3 Left | 0.247 | 0.5115 | 0.629 | 1.280 | 0.469 – 3.493 |
| PIP 4 Left (reference) | 0□ | — | — | — | — |
| **Interaction Terms** |  |  |  |  |  |
| FFI × PIP 2 Right | 0.019 | 0.0161 | 0.248 | 1.019 | 0.987 – 1.051 |
| FFI × PIP 3 Right | 0.026 | 0.0195 | 0.181 | 1.026 | 0.988 – 1.066 |
| FFI × PIP 4 Right | 0.006 | 0.0158 | 0.720 | 1.006 | 0.975 – 1.037 |
| FFI × PIP 2 Left | -0.023 | 0.0152 | 0.125 | 0.977 | 0.948 – 1.007 |
| FFI × PIP 3 Left | 0.000 | 0.0176 | 0.990 | 1.000 | 0.966 – 1.035 |
| FFI × PIP 4 Left<br>(reference) | 0□ | — | — | — | — |

In the model, no significant interaction effects were observed between FFI and PIP joint number, indicating that the association between FFI and swelling did not differ significantly across joints. The interaction terms for FFI × PIP joint were all non-significant (all p > .05), with effect estimates close to null (Exp(B) ranging from 0.977 to 1.026), suggesting minimal evidence of effect modification by PIP joint number.

The model indicated a statistically significant negative association between mean FFI and swelling (B = −0.056, SE = 0.0185, p = 0.002; Exp(B) = 0.945, 95% CI: 0.912–0.980), suggesting that higher mean FFI was associated with lower odds of swelling. In contrast, the visit and joint specific FFI effect was not statistically significant (p = 0.519; Exp(B) = 1.009, 95% CI [0.982–1.037]), indicating no evidence of an immediate association between FFI and swelling.

#### Longitudinal correlation between changes in joint-specific FFI and clinical PIP joint swelling

We assessed the correlation between changes in PIP joint-specific FFI and changes in PIP joint-specific swelling between two visits using Spearman’s rho for PIP joints 2–4 of both hands separately (Table 3). Changes in FFI and changes in PIP joint swelling showed only weak correlations (rho = −0.005 to 0.357). While the p-values for PIP 3 left and PIP 2 right suggest evidence of an association, the CIs included the null value and therefore the result should be interpreted with caution.

**Table 3.**
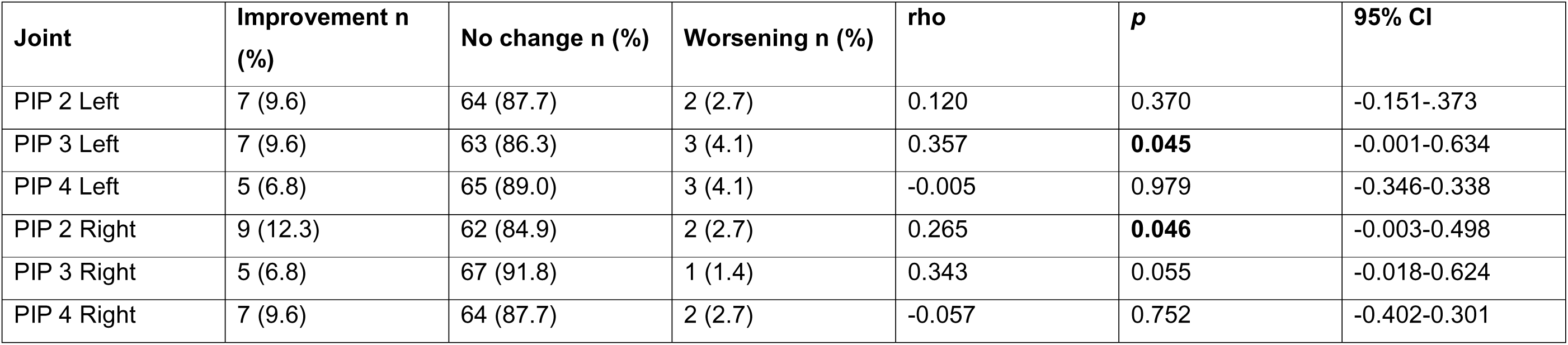
Changes of PIP joint swelling between baseline and follow-up visit and correlations between delta FFI and delta swelling.

| **Joint** | **Improvement n (%)** | **No change n (%)** | **Worsening n (%)** | **rho** | ***p*** | **95% CI** |
| --- | --- | --- | --- | --- | --- | --- |
| PIP 2 Left | 7 (9.6) | 64 (87.7) | 2 (2.7) | 0.120 | 0.370 | -0.151-.373 |
| PIP 3 Left | 7 (9.6) | 63 (86.3) | 3 (4.1) | 0.357 | **0.045** | -0.001-0.634 |
| PIP 4 Left | 5 (6.8) | 65 (89.0) | 3 (4.1) | -0.005 | 0.979 | -0.346-0.338 |
| PIP 2 Right | 9 (12.3) | 62 (84.9) | 2 (2.7) | 0.265 | **0.046** | -0.003-0.498 |
| PIP 3 Right | 5 (6.8) | 67 (91.8) | 1 (1.4) | 0.343 | 0.055 | -0.018-0.624 |
| PIP 4 Right | 7 (9.6) | 64 (87.7) | 2 (2.7) | -0.057 | 0.752 | -0.402-0.301 |

Overall, 40 joints improved from baseline to follow-up (swollen to non-swollen), whereas 13 joints developed new swelling (Table 3). All remaining joints showed no change in swelling status. Among the 13 joints with incident swelling, FFI values were missing at either baseline or follow-up for five joints, precluding calculation of delta FFI. Of the remaining eight joints, seven (87.5%) showed a positive delta FFI consistent with the change in swelling status, whereas one (12.5%) showed a delta FFI in the opposite direction. Among the 40 joints in which swelling resolved, delta FFI could not be calculated for 15 joints because of missing FFI values. Of the remaining 25 joints, the delta FFI correctly reflected the change in swelling status in 17 (68.0%) joints, whereas it was inconsistent in 8 (32.0%).

Representative examples of concordant and discordant changes between delta FFI and clinical swelling are presented in Figure 4.

**Figure 4.**
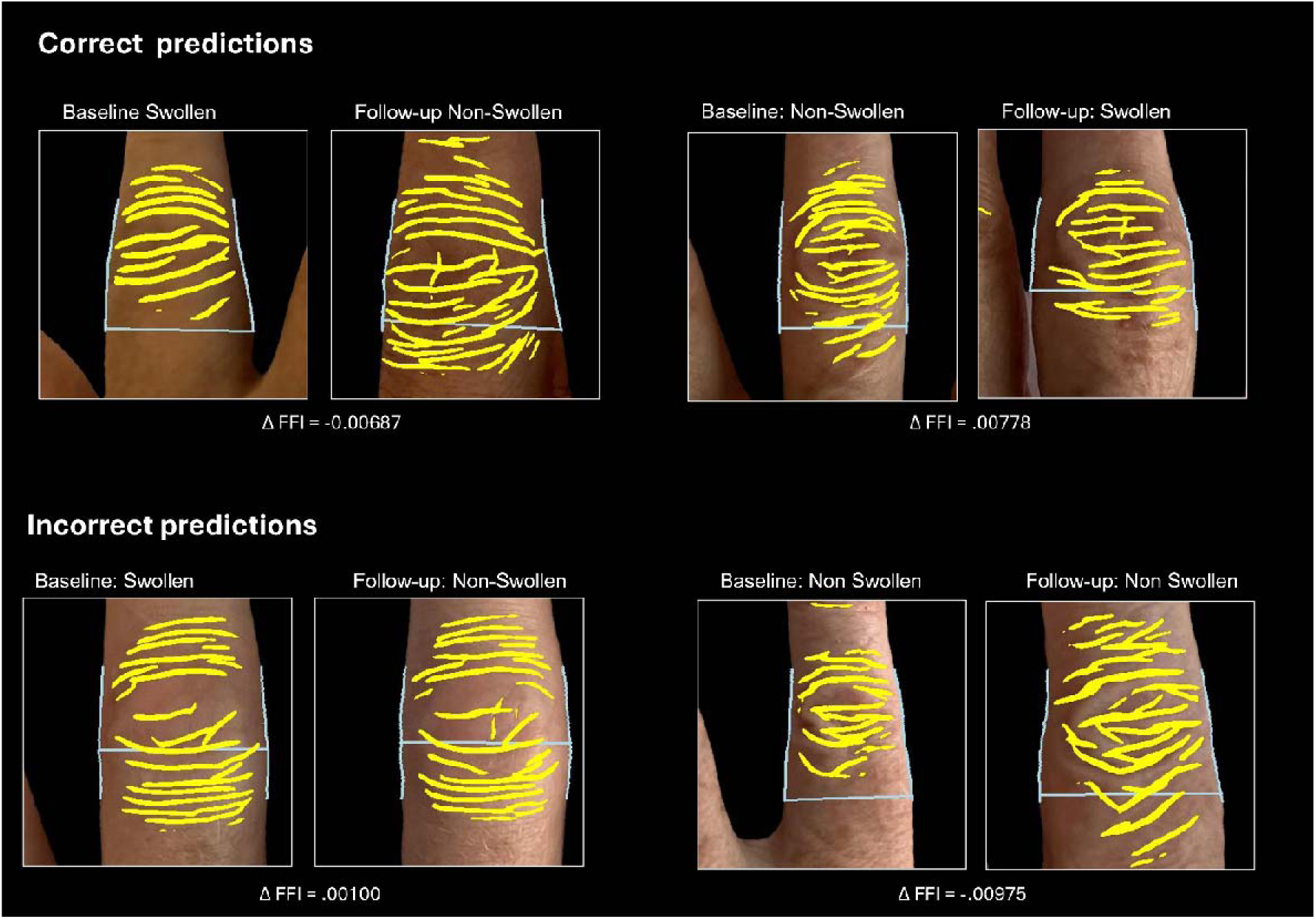
Representative examples of correct and incorrect prediction of changes in joint swelling by delta finger fold index (ΔFFI). The upper panel shows examples in which the direction of ΔFFI correctly reflected the clinical change in joint swelling from baseline to follow-up (resolution of swelling or development of new swelling). The lower panel shows examples in which the direction of ΔFFI was inconsistent with the observed clinical change. Yellow lines indicate the automatically detected finger fold patterns, joint diameters are shown in grey, and the corresponding ΔFFI values are shown below each image pair.

### Patient collected image analysis

#### Correlation between the mean FFI with the RADAI-5 score of the RA cohort

159 participants contributed images via the mySCQM app. Images provided during the duration of study ranged between 1 and 39. Usual frequency for answering PROs in the SCQM registry is once per month. 61.6 % (98/159) of mySCQM users were diagnosed with RA and filled therefore additionally PROs. They were predominantly female 72.4 % with a median disease duration of 12 years and a mean RADAI-5 score of 3.167 (Table 1, supplementary material). Due to multicollinearity of the components of the RADAI-5 score, we only correlated the RADAI-5 with the mean FFI. Mean FFI did not correlate with the RADAI-5 score (r = 0.007, *p* = 0.932, 95% CI [-0.169-0.183]).

#### Comparison of numbers and quality of images collected by HCP vs. collected by patients

The proportion of images retained for analysis was similar between HCP- and patient-collected images, with retention rates of 73.5% and 78.3%, respectively. Patients contributed over the time of study on average 7 images and HCPs collected on average 3.4 images per patient. Due to an insufficient number of images taken the same day by patients and HCPs, we could not evaluate the agreement of the FFI derived from images acquired by patients and HCPs.

## Discussion

In this study, we evaluated a machine learning–based digital biomarker derived from PIP joint photographs in a real-world cohort of patients with RA and PsA. In addition to investigating the association between the finger fold pattern and clinical disease activity, we compared the performance of the algorithm pipeline and image quality between patient-collected smartphone photographs and images acquired by HCPs.

A major finding of this study is the high feasibility of smartphone-based hand image acquisition in both patients and HCPs. Image capture was well accepted in routine clinical practice as well as through the mySCQM mobile application. Image quality was comparable between HCP- and patient-collected photographs, with 73.5% and 78.3% of images, respectively, considered suitable for analysis. Furthermore, patients participating through the mySCQM app remained engaged throughout the study period and, in absolute numbers, contributed more images than HCPs, with an average of seven hand photographs per patient. These findings demonstrate that patients with RA and PsA are able to acquire standardized hand photographs despite pain, impaired hand function, or joint deformities. This supports the feasibility of integrating patient self-collected hand photographs into remote monitoring strategies between routine clinical visits.

The principal clinical finding was the weak but statistically significant correlation between mean FFI and DAS28-CRP. Since the DAS28-CRP includes swollen joint count as one of its components, this association suggests that the FFI may capture clinically relevant manifestations of inflammatory disease activity. However, DAS28-CRP is a composite disease activity measure that also reflects tender joints, patient global assessment, and systemic inflammation through CRP. Consequently, the observed association cannot be attributed exclusively to joint swelling, and longitudinal changes in DAS28-CRP are not necessarily clinically meaningful, particularly in patients with low disease activity.

At the individual joint level, we did not observe significant correlations between longitudinal changes in joint-specific FFI and changes in clinically assessed PIP joint swelling. Nevertheless, the ability of the FFI to identify changes in swelling was encouraging. Among joints in which swelling resolved, 68.0% of evaluable cases were correctly classified by the delta FFI, whereas 87.5% of joints developing incident swelling were correctly identified. Although the number of joints with clinical changes was limited, reflecting the low disease activity of this real-world cohort, these findings suggest that the FFI is capable of detecting clinically relevant changes in joint swelling in a substantial proportion of cases.

The absence of significant longitudinal associations in the GLMM should therefore be interpreted cautiously. Several cohort-specific factors may have contributed to the lack of significant longitudinal associations. Compared with our previous cross-sectional study, no graded swelling scores (0–3) were available, as joint swelling was assessed during routine clinical visits. We have previously shown that the FFI performs best in joints with pronounced swelling [17], which may partly explain the lack of significant longitudinal associations observed in the present analysis. Furthermore, the cohort consisted predominantly of patients in low disease activity or remission, resulting in few swollen joints and only limited changes in PIP joint swelling between visits (Table 3). Consequently, only 13 joints developed incident swelling during follow-up, of which only eight were evaluable because of missing FFI data, substantially limiting the statistical power to detect longitudinal associations.

Despite standardized image acquisition instructions, 10.16% of images were excluded prior to the algorithm analysis. At present, this image quality assessment has been performed manually, a process that is time-consuming and limits scalability. These findings highlight the need for technology-assisted image acquisition guidance to reduce poor-quality image input. Improving image quality is essential for establishing an automated smartphone-based assessment of joint swelling using the FFI, which could facilitate remote monitoring while reducing the time and resources required for clinical evaluation.

Following image processing by the ML algorithm, 23.31% of individual PIP joint images were excluded due to misclassification, including cropping errors, missing or incorrectly detected joint diameters, skin folds, and other image artefacts (Figure 3). These findings indicate that the current FFI algorithm still requires further refinement. Future algorithm development should focus on improving the detection and exclusion of image artefacts through additional machine learning training, thereby increasing robustness under real-world conditions. In addition, external factors such as lighting conditions, camera angle, smartphone type, skin tone, and hand positioning should be systematically evaluated, as they may influence algorithm performance.

The resulting missing FFI values reduced statistical power and limited our ability to construct sufficiently dense longitudinal datasets for evaluating within-joint changes over time. At present, image quality assessment is still performed manually, making the analysis time-consuming and limiting scalability. Automated image quality control and real-time acquisition guidance within the smartphone application could substantially reduce poor-quality image input and facilitate reliable fully automated assessment of joint swelling in future remote monitoring applications.

Another important limitation is the lack of an imaging-based reference standard. Clinical assessment of joint swelling served as the reference in this study; however, clinical examination has well-recognized limitations. Compared with MRI or US, clinical examination demonstrates lower sensitivity for detecting synovitis [18], [19]. Furthermore, imaging studies have shown that subclinical inflammation is frequently present in patients considered to be in remission according to clinical disease activity scores and in joints classified as non-swollen on clinical examination [20], [21]. Future validation studies should therefore compare FFI-derived measures with ultrasound or MRI rather than relying exclusively on clinical examination.

External validation also remains necessary before clinical implementation. Nevertheless, our previous monocentric study demonstrated a similar association between FFI and joint swelling, supporting the reproducibility of the underlying concept [17].

The exclusive analysis of PIP joints represents another limitation. Disease activity in RA and PsA also frequently affects the metacarpophalangeal (MCP) joints, and algorithms for automated MCP joint assessment are currently under development. Combining information from multiple joint groups may further improve diagnostic performance.

Finally, our exploratory analyses demonstrated no correlation between FFI and the RADAI-5. This finding is not unexpected, as the RADAI-5 assesses patient-reported disease activity, pain, and morning stiffness rather than objective joint swelling. Previous studies have similarly demonstrated that painful and swollen joints only partially overlap [22]. Moreover, no information on patient self-assessment of swollen joints was available, preventing direct comparison between patient-reported swelling and FFI-derived measures. Consequently, the FFI may be most informative when combined with complementary patient-reported outcome measures rather than being used as a standalone indicator of overall disease activity.

Finally, the lack of an imaging-based reference standard represents an additional limitation. Although clinical assessments of PIP joint swelling were available, no imaging data on synovitis were collected to allow comparison with FFI-derived measures. While clinical assessments of PIP joint swelling were available, no imaging data for synovitis were available to allow correlation with the FFI. Studies have shown that clinical assessment is limited, as subclinical inflammation is often present in patients who are in remission according to disease activity scores, as well as in joints that are assessed as unaffected by clinical examination

Despite these limitations, one of the major strengths of this study is the evaluation of smartphone-based hand imaging in a real-world setting. Both patients and HCPs showed high acceptance of image acquisition, demonstrating the practical feasibility of integrating this technology into routine care and remote monitoring.

In conclusion, this longitudinal study demonstrates that patient self-collected smartphone hand photographs can be successfully integrated into an established mHealth platform with sustained patient engagement. Although further improvements in image acquisition support, automated quality assessment, and algorithm performance are required, these findings represent an important step toward reliable, automated smartphone-based monitoring of inflammatory joint swelling in patients with RA and PsA.

## Author contribution

CNK: Project management, methodology, investigation, data curation, statistical analysis, visualization, writing and revision of the manuscript.

JM: Data curation, software and revision of the manuscript.

CP: Data curation, and revision of the manuscript.

MB: Conceptualization, methodology, revision of the manuscript.

AD, DD, LB, MN, MA, AS, ARR, CIK, TM, BM, CM: Investigation and revision of the manuscript.

JG: Statistical analysis, validation and revision of the manuscript.

TH: Conceptualization, project management, resources, funding acquisition, investigation, methodology, visualization, writing and revision of the manuscript, supervision.

## Data availability

This study is based on data collected by the SCQM Foundation (Swiss Clinical Quality Management in Rheumatic Diseases), which operates a nationwide registry for rheumatic diseases. Access to the data is subject to restrictions and requires approval from the SCQM Foundation in accordance with the SCQM Rules of Research and Collaboration (https://www.scqm.ch/en/research/research-with-scqm-data/). Interested parties may contact the SCQM Foundation to request access to the data for research purposes.

## Acknowledgments

We would like to thank the SCQM registry and participants of the study. A list of rheumatology offices and hospitals contributing to the SCQM registry can be found on https://www.scqm.ch/en/aboutscqm/active-institutions/.

## Conflict of interests

CNK, JM, JG, none declared. MB & TH: are shareholders of Atreon SA. TH has filed a patent jointly with CHUV related to the image analysis for the monitoring of swelling of body parts

## Funding

This study has received an unrestricted grant from Fresenius Kabi.

## Notes

### Author Declarations

The study was approved by the Commission cantonale d'ethique de la recherche sur l'etre humain (CER-VD) (2020-00033) and conducted in accordance with the Declaration of Helsinki.

