## Supplementary Material for "Smartphone Imaging for Remote Monitoring of Inflammatory Arthritis in a Real-World Cohort: Longitudinal Evaluation of a Machine Learning-Based Finger Fold Biomarker"

**Table 1. Patient characteristics and PROs of mySCQM users diagnosed with RA.**

| Variables | RA Cohort, N = 98 |
|---|---|
| Age, mean (SD) | 56.6 (13.639) |
| Female, n (%) | 71 (72.4) |
| Disease duration, median (IQR) | 12 (15) |
| RADAI-5, mean (SD) | 3.167 (2.2483) |
| Pain level today, median (IQR) | 2 (4) |
| Global patient estimate disease activity, median (IQR) | 2 (4) |
| Activity of rheumatic disease today, median (IQR) | 2 (4) |
| Activity of rheumatic disease last 6 months, median (IQR) | 4 (4) |
| *Normally distributed variables are described with mean (SD), non-normally distributed variables with median (IQR). Missing values for the RADAI-5 were N = 12 and for disease duration N = 4.* | |
